# What do we have here? A Systematic Review of Mental Health Policy in Colombia

**DOI:** 10.1101/2024.03.11.24304116

**Authors:** Norha Vera San Juan, German Alarcón Garavito, Monica Gonzalez Gort, Maria Cecilia Dedios Sanguinetti, Rochelle Burgess, Diego Lucumí

## Abstract

**Background:** Colombia’s mental health policy is currently under the spotlight due to the global call of prioritising mental health services quality and innovation and the Colombian vision of having an articulated mental health system based on inclusion and community participation. Despite the interest in this topic and proliferation of policy documents, there is no clarity around mental health concepts underlying the compendium of mental health policies, plans and legislation, which are crucial for successful development and implementation of public mental health.

**Method:** This study is a novel Systematic Collaborative Policy Review which includes a structured approach to identifying and synthesising relevant institutional documents, alongside a realist approach including consultations with experts throughout the review process to increase the applicability of results.

**Results:** 295 records were screened at title and main content stage, 66 were assessed in full text and 46 were included in this review. Most documents identified were created by the Ministry of Health, the Unit for the Attention and Integral Reparation to the Victims, and regional governments.

We found Colombian institutional documents had a holistic understanding of mental health, including considerations around the importance of prevention through the creation of healthy protective environments, as well as protecting right to receive a diagnosis, treatment and rehabilitation for people who require mental health care. There was a strong focus on childhood wellbeing and addressing issues related to illegal drug use. Numerous preventive programmes were listed such as specific programmes for LGTB+ and indigenous communities.

**Conclusion:** Colombia has a strong mental health legal and programmatic baseline, with ample coverage of aspects ranging from prevention and promotion to quality treatment. However, the extent to which these plans are implemented is unclear. There is a need for clear implementation paths to be included in policy plans, and identification of measurable outputs to monitor success.

## Introduction

Colombia’s mental health services and policy are currently under the spotlight due to the global call of prioritising mental health services quality and innovation, as well as the Colombian vision of having an articulated mental health system based on inclusion and community participation (1). To measure changes and trends in mental health necessities, three national studies about mental health in Colombia were conducted in 1993, 2003 and 2015. These studies have been vital to inform mental health policy, plan, and legislation, identifying the primary mental health problems, and linking these to social crises such as the armed conflict or economic downturns (2,3).

The most recent nationally representative study in 2015 surveyed 13.555 households and 15.351 people, including children between 7-17 years old. The study reported four critical areas of focus for public mental health: psychoactive substance use, suicidal conduct, violence and conviviality (*Convivencia in Spanish*). In response, related policies and laws were designed to address these areas. Essential mental health-specific policy documents to date are the Mental Health Law (MHL) (1616/2013); Resolution 4886 (2018), that introduced the National Mental Health Policy (NMHP); and the CONPES 3992 (*Consejo Nacional de Política Económica y Social in Spanish)* public policy roadmap about strategies to promote mental health in Colombia. However, numerous complementary institutional documents lay out the strategy to promote wellbeing and address mental health problems in Colombia, ranging from documents developed for national purposes, for specific regions, and also developed by different ministries and public institutions.

Healthcare policy in Colombia is predominantly governed and overseen by the Ministry of Health and Social Protection (MinSalud). However, various public and private organisations actively contribute to formulating intersectoral strategies for the national health agenda, and this is established as a political priority in the CONPES 3992. Stakeholders range from other government institutions, non-governmental organisations (NGOs), international health organisations, private healthcare providers, and academia. Some key government institutions are ministries, the National Planning Department (DNP), The National Administrative Department of Statistics (DANE), and The Colombian Family Welfare Institute (ICBF). The latter actors are mainly involved in diagnosis, planning, and funding allocation, however, implementation and evaluation primarily rely on regional and local entities, such as municipal health secretaries (4).

Despite the interest in this topic and proliferation of institutional documents in the country (5,6), there is no clear understanding of the definitions and structures that support Colombian mental health policy, plans and legislation. Public mental health plans in Colombia to date have evolved organically, mainly based on individual policy-makers’ knowledge or mirroring foreign systems, rather than based on comprehensive mental health models, stakeholder participation, and local evidence. A thorough review and evaluation of the institutional documents that secure mental wellbeing in Colombia has not been conducted to date, there is no certainty around the coherence across documents produced by different institutions, or the quality of national policies and plans in relation to international standards. This information is crucial for successful mental health system development and implementation.

Concurrently, governments make great efforts to generate fair and evidence-based policies, however, the research methods we found available did not allow for generating timely rigorous results, and mental health system actors (stakeholders) were not being included in developing or monitoring the implementation of policies, plans and laws. This has impact in more rural contexts and/or territories affected by violence, poverty, illicit economies and institutional weakness (called PDET territories in Colombia, based on the plan Development Programs with Territorial Focus, to foster their development), as they face challenges to implement national regulations in practice.

For the reasons listed above, we set out to conduct a review of institutional documents (policies, plans and legislations) about mental health in Colombia, with a systematic and applied approach. We aimed to (1) define mental health structures, understandings, and focuses of policy documents (2) situate these findings in practice, and (3) evaluate the quality of mental health policy considering international standards.

## Methods

Policy reviews are often conducted in ways that lack a systematic approach to identifying documents and engaging with the material. Knowledge synthesis methods used to inform policy have been found to be highly subjective and not reproducible (7). Rigorous knowledge synthesis methods that intend to cover policy-related information (8) are focused on reviewing scientific data and are not designed with efficiency and applicability in mind in order to facilitate evidence-based decision-making.

To overcome this, we developed a systematic methodology titled a Systematic Collaborative Policy Review). It can be summarised as combining relevant steps followed in systematic literature reviews (8), collaborative research approaches used in Rapid Evidence and Rapid Realist Reviews (9), and national policy and plan evaluations as recommended by the world Health Organisation (WHO) (10). In line with rapid research approaches for applied research, stakeholders including patient representatives, clinicians, and policymakers provided feedback refining the scope of the review questions, search strategy, data extraction form, interpretation of findings, and articulating applied theories. Also, a protocol was developed a priori and registered on the Open Science Framework (OSF) repository. More detail on each step of the method is provided below.

### Stakeholder consultations

The research team mapped relevant stakeholder groups based on our knowledge from previous work on mental health policy and planning in Colombia. Stakeholders included service user representatives, policymakers, clinicians, and civil servants.

Potential contributors were given information about the project and asked if willing and available to provide input based on their expertise by experience at key stages of the research process (i.e., developing the search strategy, data extraction, and interpretation of results). Consultations were carried out by GA-AG and NVSJ via email and/or videocall. Information was shared in advance in accessible formats and specific questions were included to guide the conversation to facilitate meaningful contributions from all stakeholders. A protocol was developed before searching and is registered in the Open Science Framework (OSF) platform (11).

### Search strategy

Relevant Colombian regional and national government websites were mapped and corroborated in meetings with stakeholders. We conducted searches including key terms related to mental health, services, and professionals; Colombian regions; and budget and expenditure. Websites searched included The National Department of Planning (DNP), Department of Social Prosperity, The Ministry of Health (MinSalud), and The Victims Unit, among others. Due to the scope and timeline for conducting this review, it was not possible to include individual searches for the websites of the 32 Colombian regions (*Departamentos*). We focused on the region of Caquetá as a case example due to (1) being the only region to be categorised in full as a PDET territory, and (2) there being a specific interest on behalf of local government in developing mental health infrastructure (12,13).

Due to the variability across website search engines, search words were adapted following a pre-established systematic approach. We ran exploratory searches in key databases to test and refine the strategy, monitoring the presence of key documents among search results. Additionally, the search was supplemented by hand searching and consulting stakeholders.

The search was conducted between June and December 2020. Websites were “live” databases, meaning that the number and type of results may vary depending on the consultation date. Notably, some government websites were not available overseas, requiring the use of a virtual private network (VPN) to access them.

A full description of the search strategy including the exact search dates and terms used for each website are included in Appendix 1.

### Eligibility criteria

For this work we considered policy, plan, and legislation documents, which are official documents emitted by governmental institutions that provide relevant information about policy actors, context, and priorities (45). Policy documents include laws, resolutions, public service plans, guidelines, reports, roadmaps, and agreements (14). Documents were included for data extraction if they contained any information relevant to mental health/wellbeing. Mental health was understood as being composed by the four dimensions included in the SPICE model of wellbeing (15), as depicted in Figure 1. Further inclusion and exclusion criteria are depicted in table 1.

**Figure 1.**
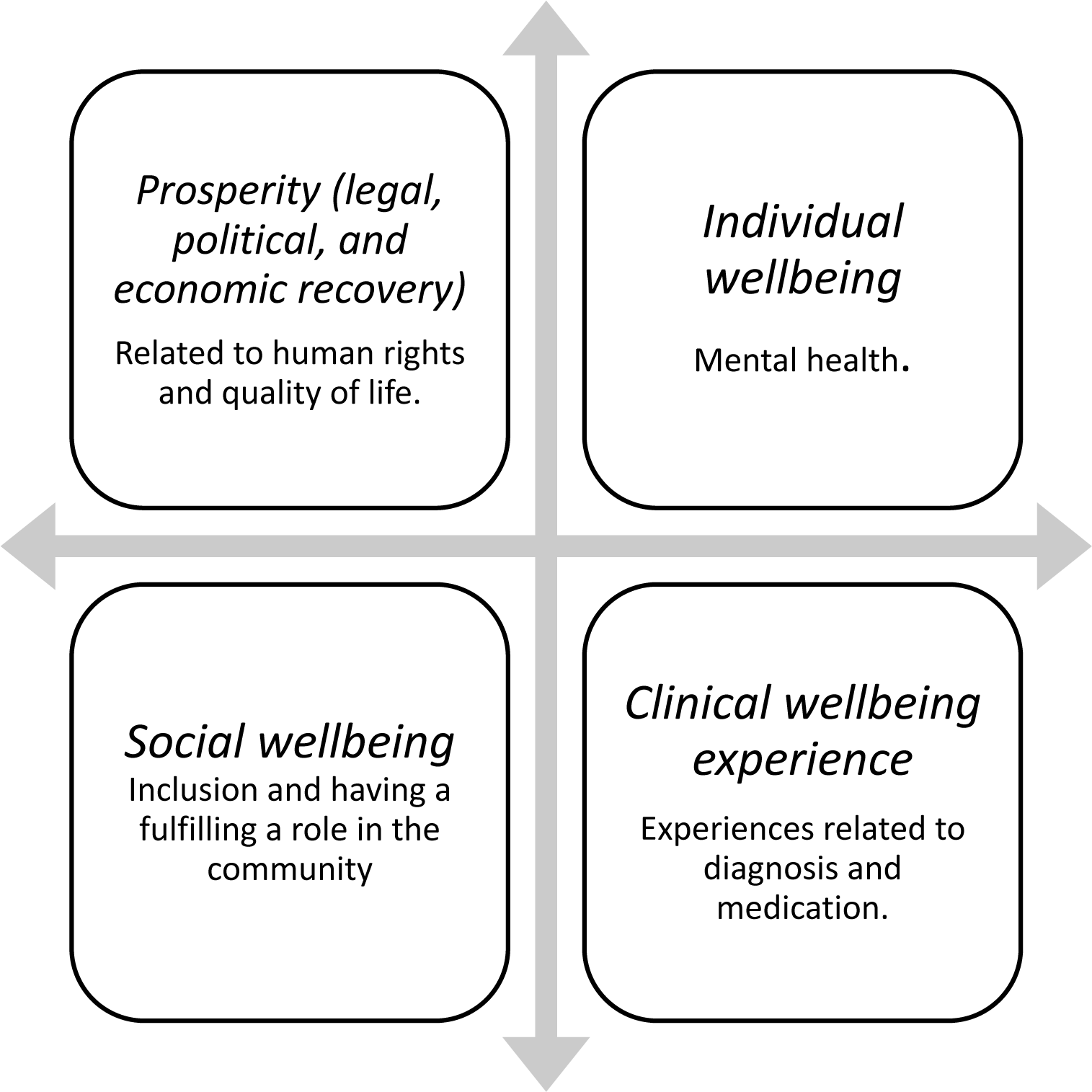
The SPICE model.

**Table 1.**
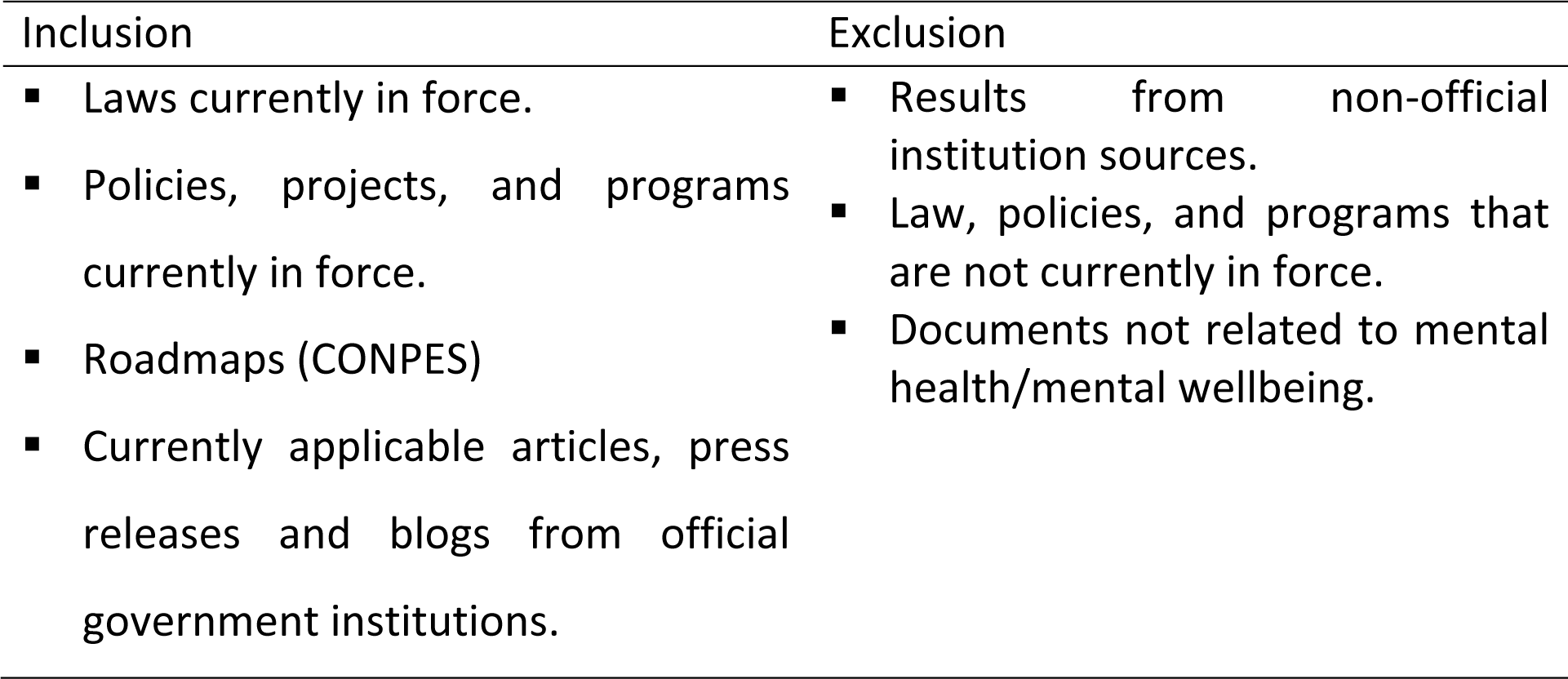
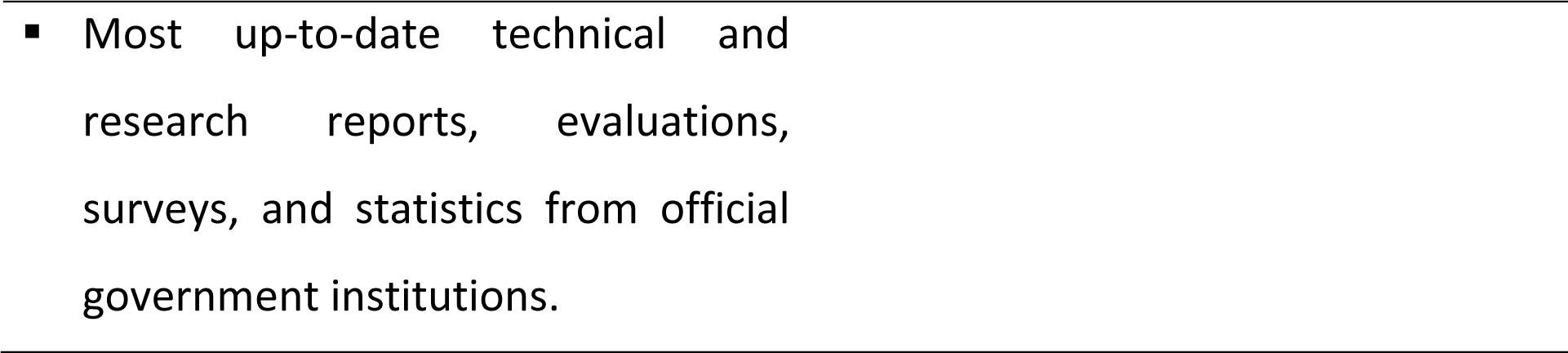
Inclusion and exclusion criteria.

Initial screening of document titles was conducted on the websites and documented on Excel. Full texts were sourced for policy documents deemed relevant, and these were screened against the full criteria, with a second author reviewing those that were not clear to establish consistency in the document selection.

### Data extraction

The data extraction form included basic information such as document title, type of document, date, region/population it is directed to, government body it was issued by, and types of mental health are covered. Additionally, the form included data extraction categories related to the National Mental Health Policy, the SPICE wellbeing model, the review research questions and aims, and key mental health topics such as substance use, suicide, the COVID-19 pandemic and government or state support offered to people with mental health problems. Stakeholders reviewed the form to ensure the data extracted was useful to cover current knowledge gaps in policy development and mental health practice.

Data extraction was conducted using a form designed on Qualtrics. The form was piloted on 10% of the included documents and was refined before extracting data from the remaining 90%. Data were extracted by two reviewers (GA-AG, MGG) and consistency checked by a third (NVSJ).

The complete data extraction form is available in Appendix 2.

### Quality appraisal

The Systematic Collaborative Policy Review process included feedback from stakeholders to reduce bias in the search and document selection process. Due to the nature of the documents being reviewed, and the collaborative method being applied, it is not considered necessary to employ a quality appraisal tool to assess each document’s veracity. The national mental health policy and plan were evaluated following WHOs checklist for evaluation a mental health policy (16). Following the checklist instructions, we scored 76 questions/statements within 20 suggested categories with a scale between one and four (1=yes/ to great degree; 2=to some extent; 3=no/not at all; and 4=unknown).

### Evidence synthesis

Due to the length of the data extracted, an appropriate big qualitative data analysis method was applied. The Collaborative and Digital Analysis of Big Qual Data in Time Sensitive Contexts (LISTEN) method connects the human insight of participatory analysis methods, and the precision and agility of digital analysis software (17). This analysis consists of iterative cycles of intercalating team discussions and the use of digital text and discourse analytics tools. Digital analysis was completed using Infranodus, a text network analysis software to measure topics, patterns, and categories occurring in extracted data (18). Manual analysis consisted in descriptive content synthesis (19) conducted by two researchers (NVSJ, GA-AG) reviewing all the data extracted and triangulating the results with the text network analysis findings. Initial synthesis of the data was kept in Spanish to maintain the same language used in institutional documents.

### Mental health policy assessment

To fulfil aim 3 of this review (evaluating the quality of Colombia’s mental health policy in light of international standards), we completed the checklist for evaluating a mental health policy by the WHO to detect content issues and evaluate overall performance (9). This checklist evaluated process issues (e.g., situation assessment, consultation, research to inform policy) and content issues (e.g., vision, principles, financing, coordination, legislation).

## Results

### Documents selection

The initial search generated 300 results across all the databases/websites. After deduplication, five documents were removed manually. The titles and summaries of the remaining 295 documents were screened by one author (GA-AG) applying the full inclusion and exclusion criteria, of which 66 documents remained for full text screening. Twenty of these were excluded, the main reasons being because they did not have meaningful content related to mental health/wellbeing, or they were no longer in vigour. All the included documents, and 25% of the excluded, were screened by second researcher (NVSJ). Forty-six policy documents were included in the final review (see Figure 2 for the Preferred Reporting Items for Systematic Reviews and Meta-Analyses (PRISMA) Flow Diagram). All of the included documents, and 25% of the excluded were double screened by another reviewer (NVSJ).

**Figure 2.**
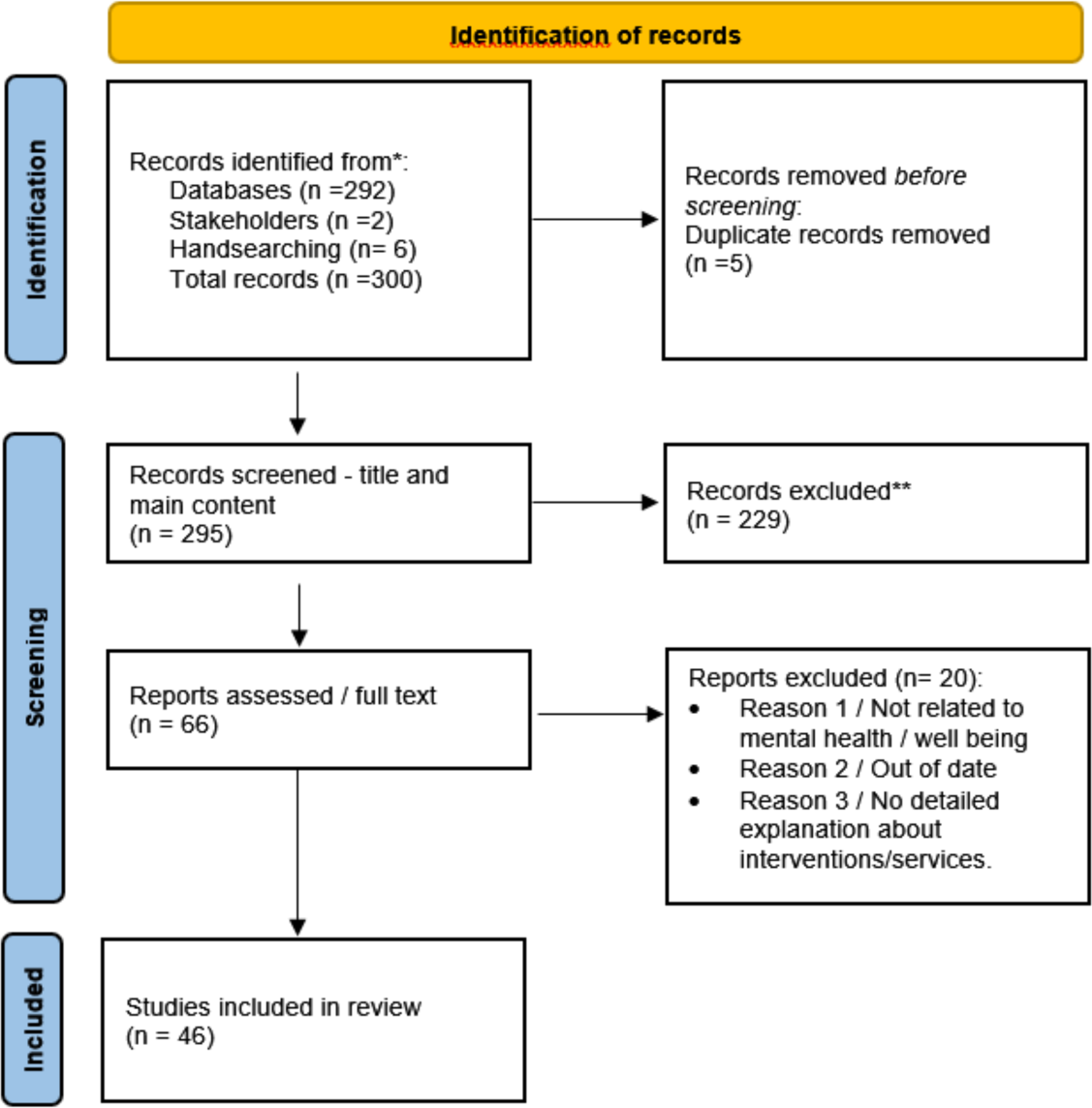
The PRISMA Flow Diagram.

### Documents characteristics

The main document characteristics are summarised in Table 2. Most documents were published by the Ministry of Health, the Unit for the Attention and Integral Reparation to the Victims, and the regional government of Caquetá. An additional five documents were suggested by the expert stakeholders. Fourteen documents were official press releases that pointed to policy documents, nine guidelines, eight government reports, and four CONPES roadmaps. Thirty-four documents were at the national level, eight at the regional (*Departamentos*) and four at the local level.

**Table 2.**
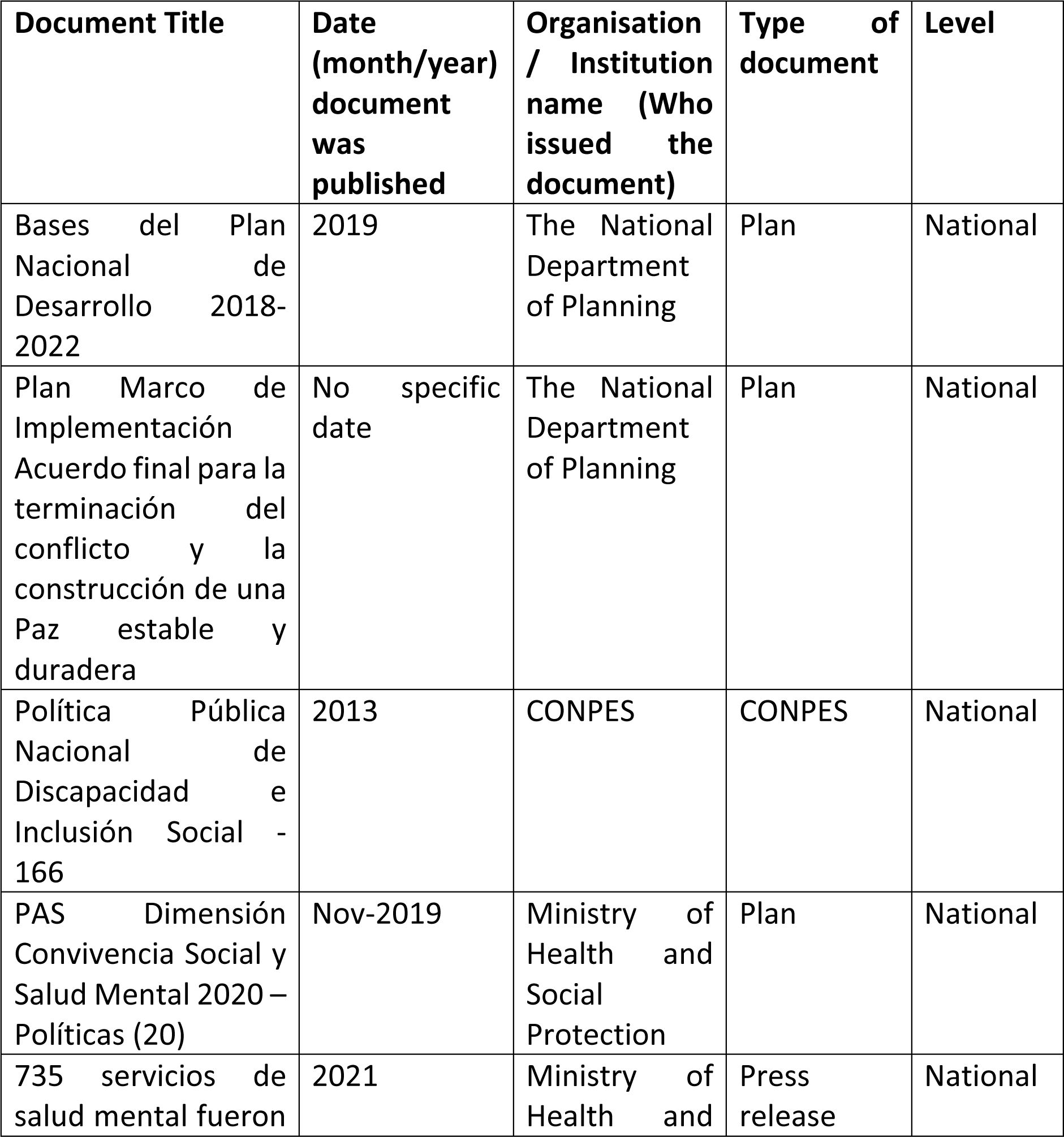
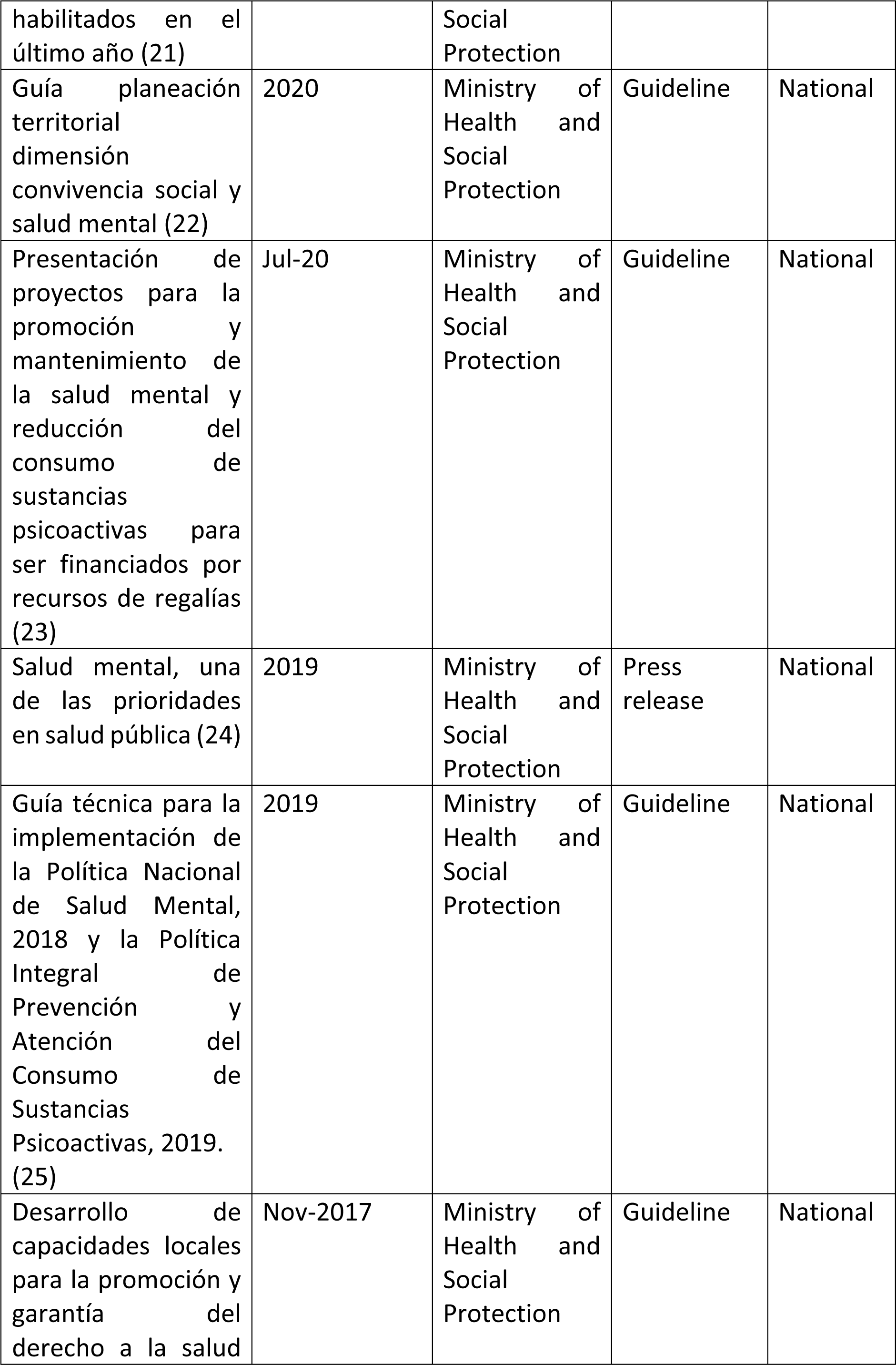
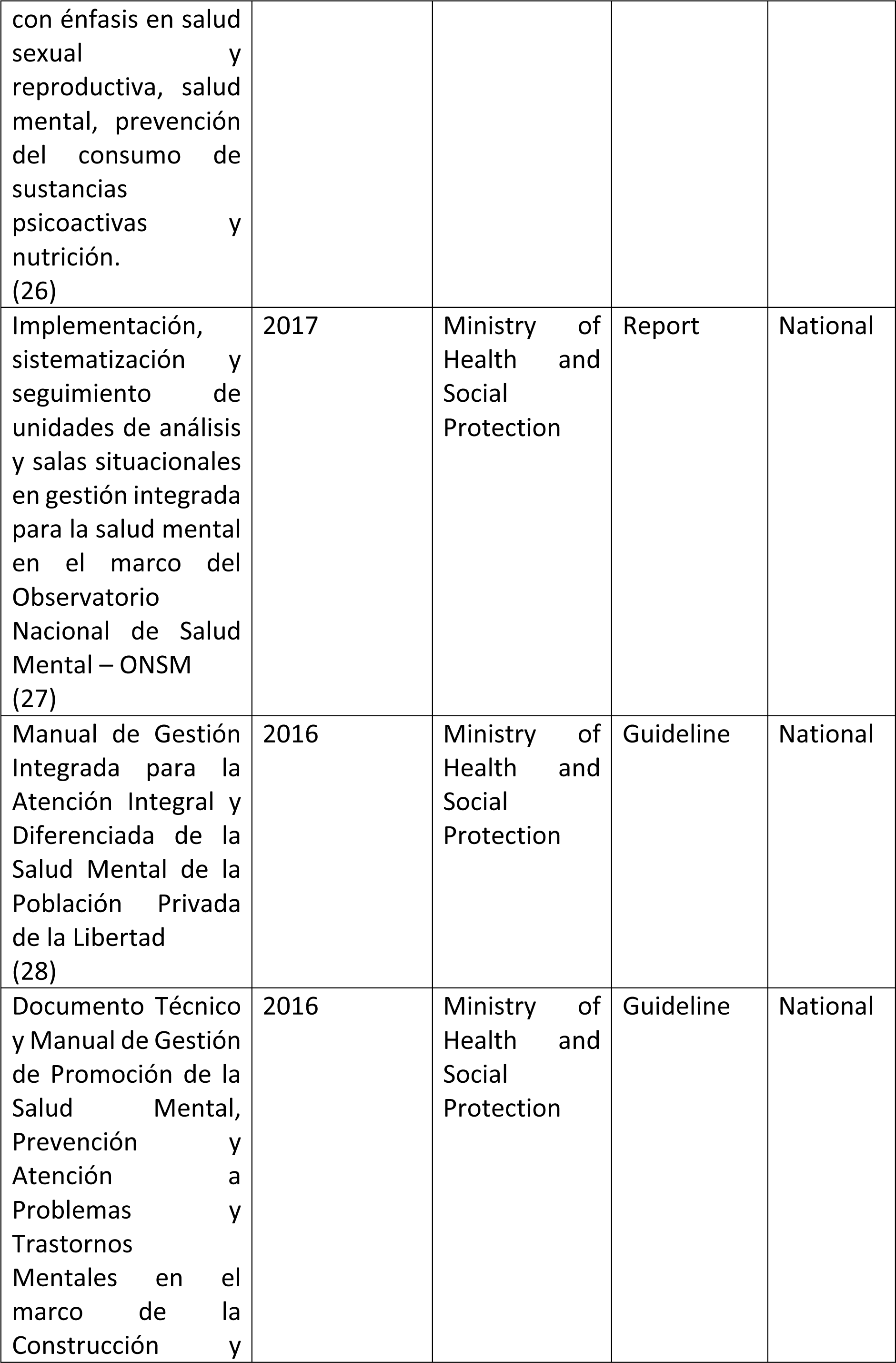
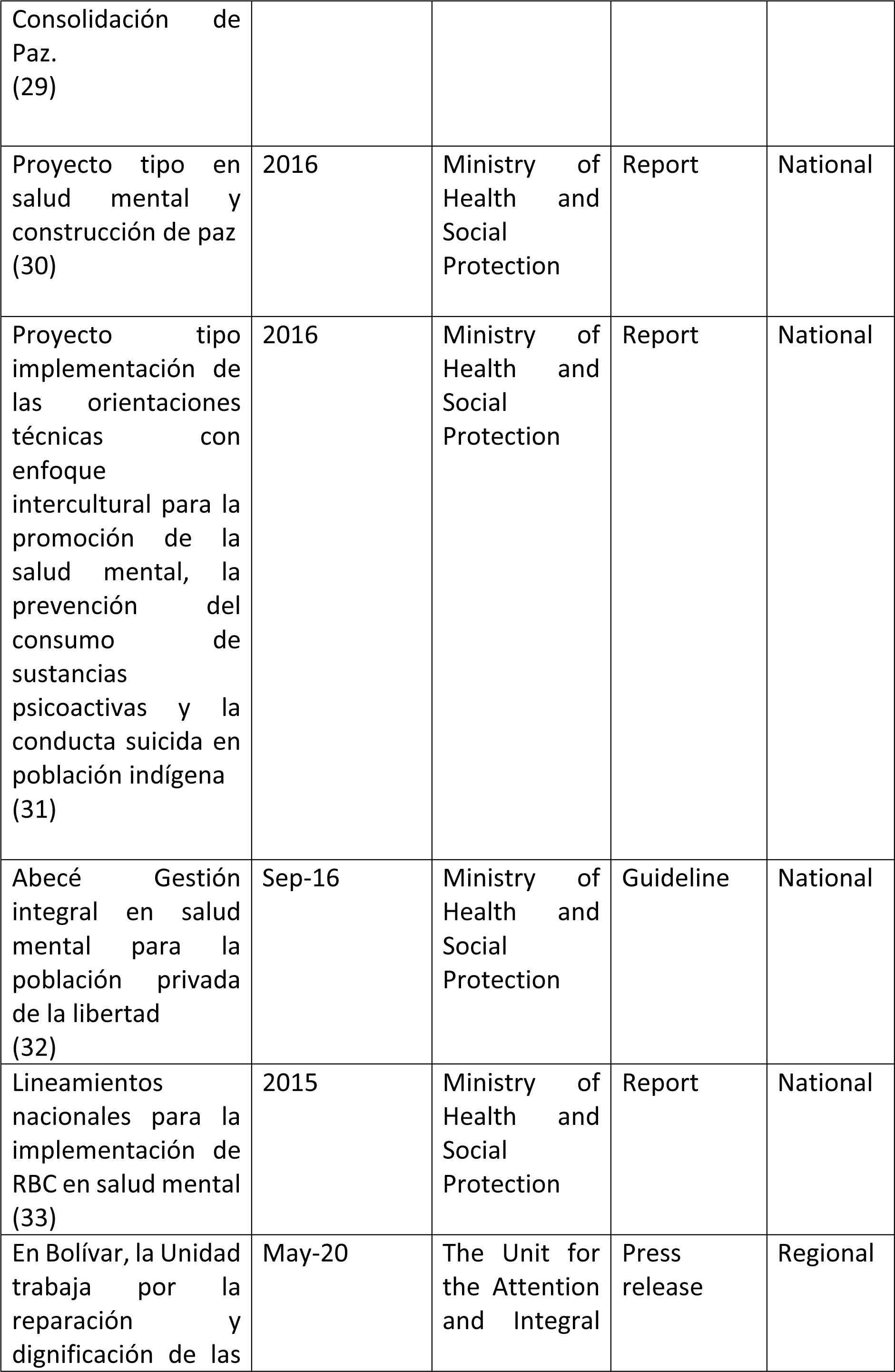
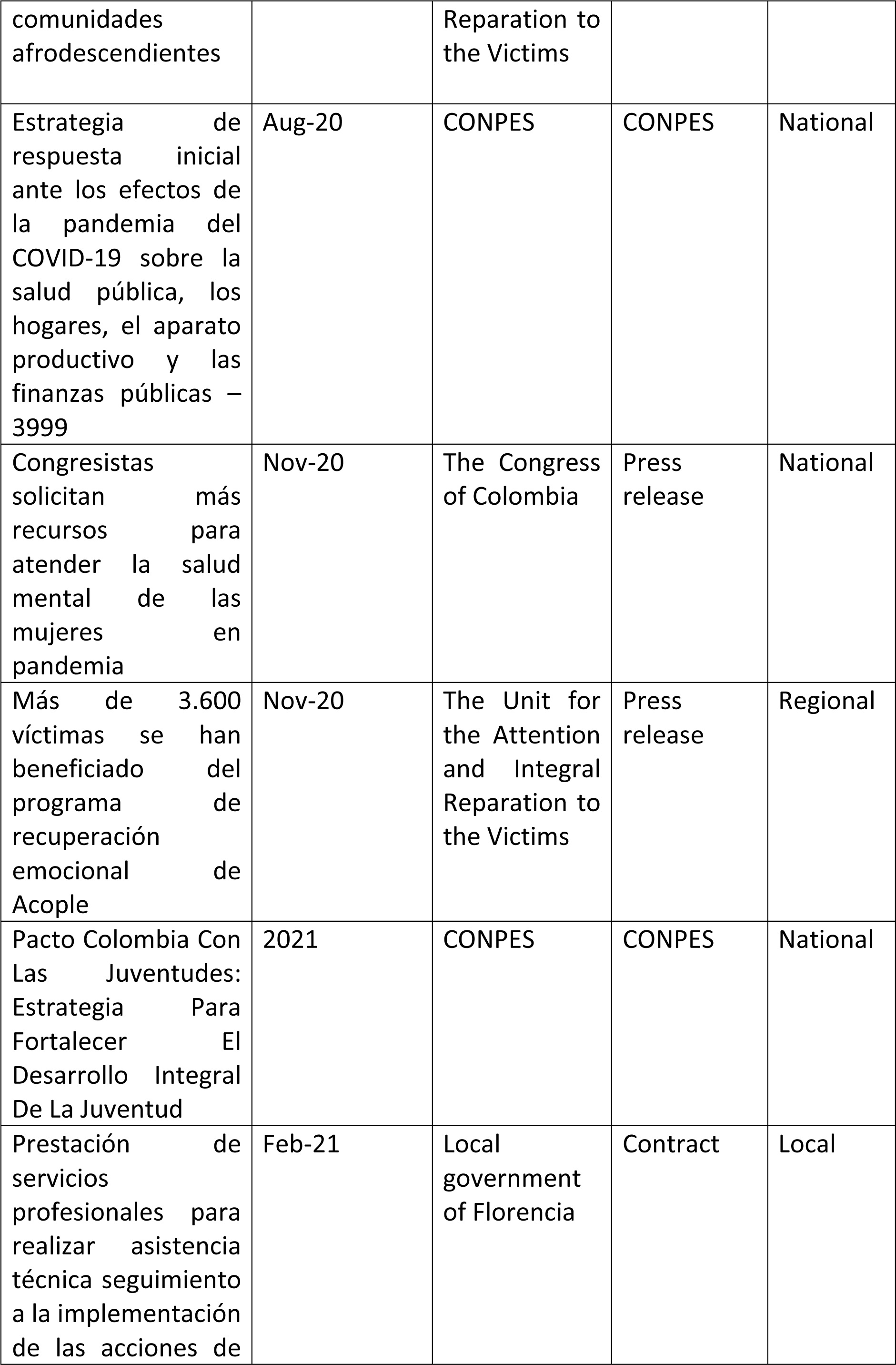
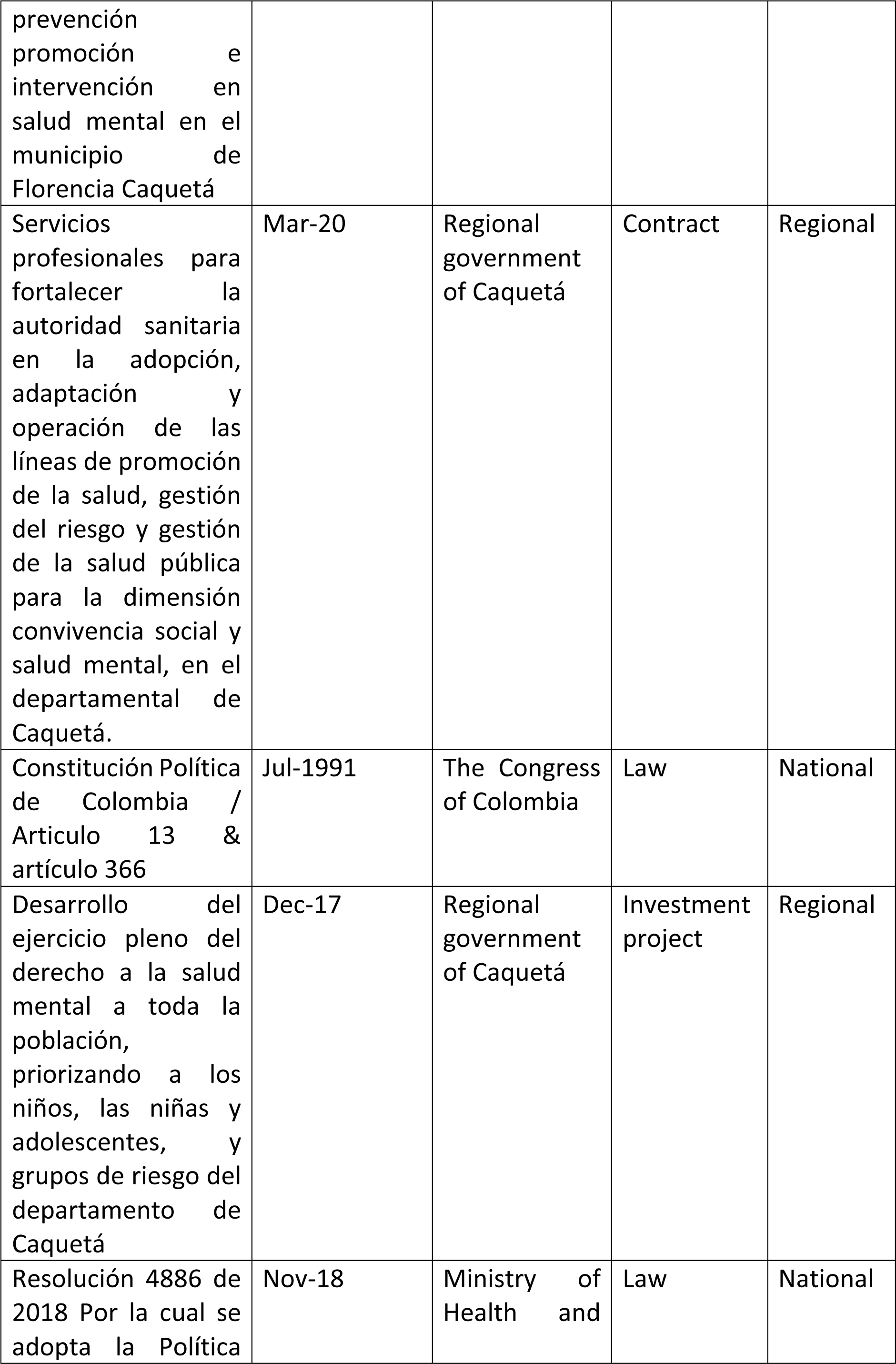
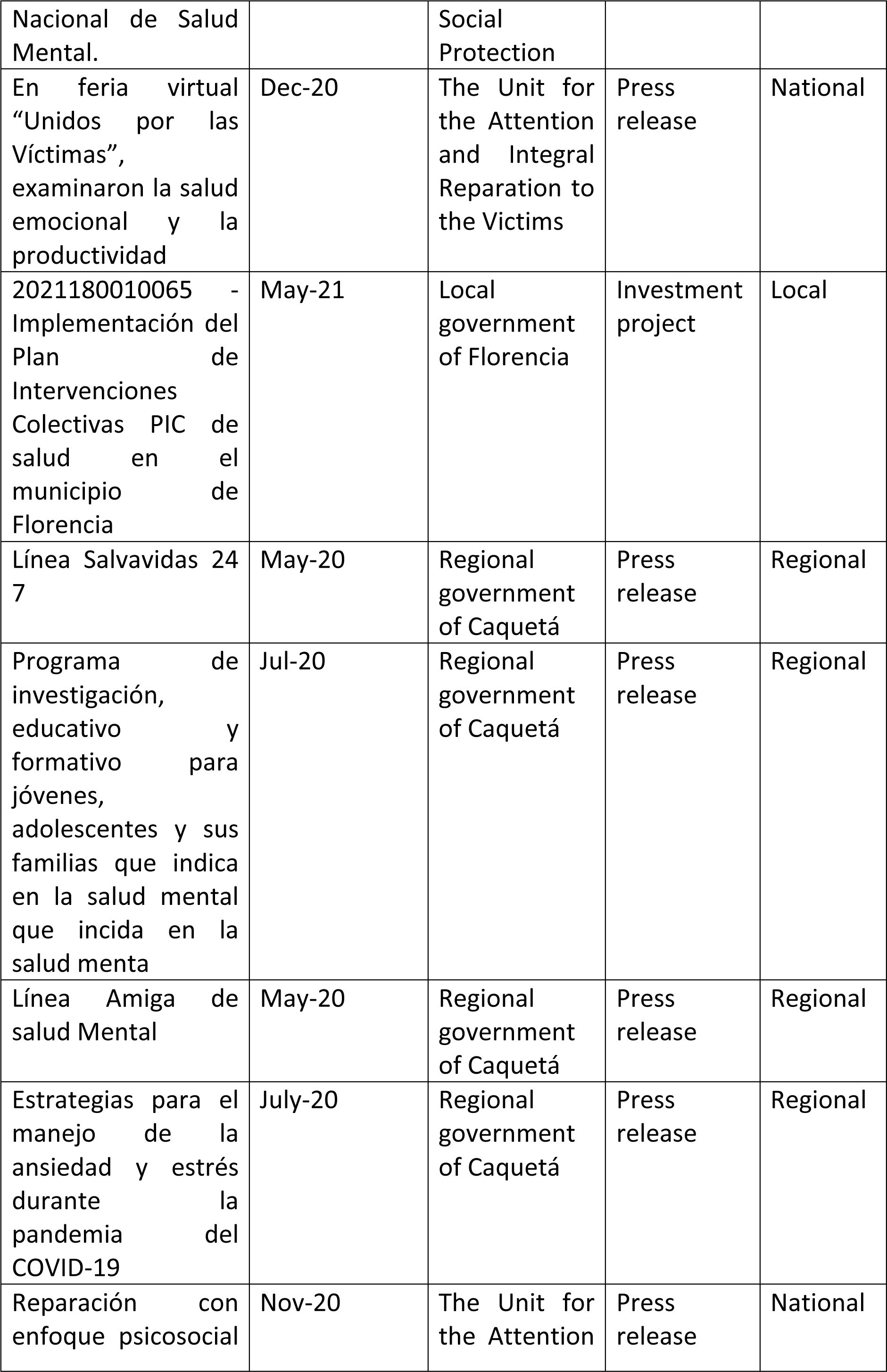
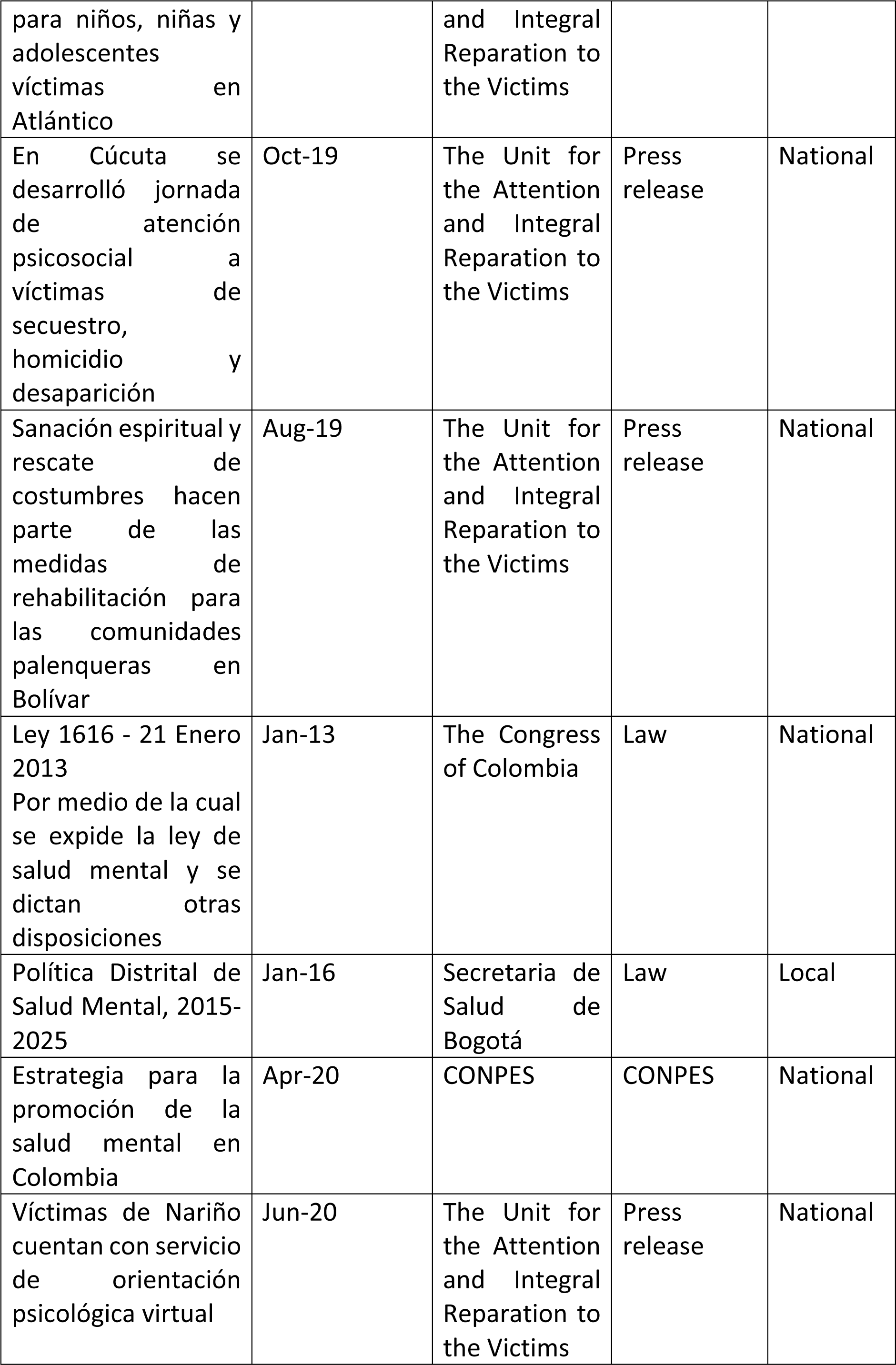
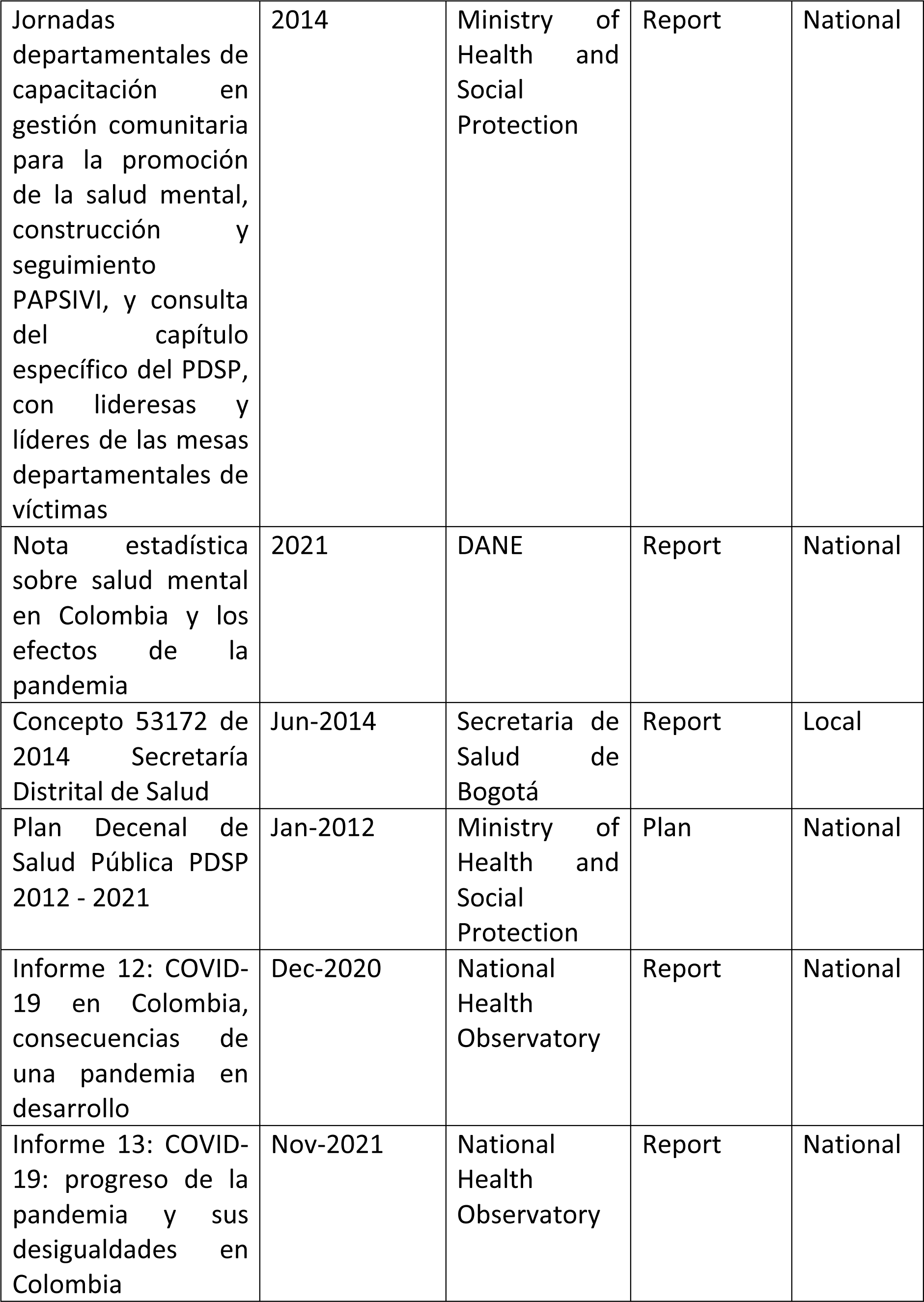
Documents characteristics.

### Sample stakeholders

We held consultations with five expert by experience stakeholders. These took place at the initial stages of planning and carrying out the search strategy, when developing the data extraction form, and to interpret results in practice. One participant worked at the Mental Health Division of the Ministry of Health, one from the National Department of Planning (*DNP*), two were activists and co-directors of a Colombian NGO seeking to dismantle psychiatric violence and centring the voice of survivors of said violence, and one was a mental health consultant and director of a private mental health service. Three of the stakeholders identified themselves as cis women, one as a cis man, and one as a trans man. Three expressed having lived experience of mental health problems and the mental health system, which they applied in their day-to-day work. Stakeholders’ input and observations are included throughout the themes that resulted from the evidence synthesis outlined below, thus situating in practice the definition of mental health structures and understandings from policy documents.

### Mental health structures and understandings in Colombian policy

This section outlines the main mental health policy documents in Colombia, the definition of mental health in Colombian policy documents, proposed determinants of mental health, and the structures intended to support mental health in the country. We then go on to synthesise specific aspects emerging in the analysis as key focus in policy documents, these are early childhood and education; cultural components; mental health promotion and psychosocial rehabilitation; and community-based rehabilitation

### Core mental health policy documents in Colombia

Colombia’s National Mental Health Law aims to “*guarantee the realisation of the right to mental health, especially in children and young people*” (1). It emphasises the obligation to provide mental health care, facilitate inclusion and guarantee people’s Constitutional rights regardless of their mental health condition. Similarly, Resolution 4886 was designed to endorse the adoption of the NMHP. To achieve this, the policy was divided into 5 thematic axes: (1) Promotion of coexistence and mental health in environments; (2) Prevention of individual and collective mental health problems; (3) Comprehensive treatment.; (4) Comprehensive rehabilitation and social inclusion; (5) Sectoral and intersectoral management, articulation, and coordination.

The NMHP has five guiding areas of focus that shape its execution, these are: human rights, life course, gender, psychosocial, and distinctions based on population and territorial characteristics.

In terms of monitoring and impact evaluation, the NMHP implementation has been assessed using indicators specified in the Ten-Year Public Health 2012-2021 (*Plan Decenal de Salud Publica in Spanish*), such as reducing violence rates, implementing monitoring mental health systems, and increasing resilience rates. For instance, available data from the National Mental Health Observatory and the *Sispro* (Colombian healthcare information system) have revealed minor variations in the mortality rate due to self-inflicted injuries (from 5.12 in 2009 to 5.29 in 2021) (34). This is an indicator used in both health policies and in charge of regional and local mental health committees (35).

Lastly, the mental health CONPES is a public policy roadmap designed by the DNP, implemented by the government’s executive power and out of the legislative power discussion. The CONPES established an analysis of the current mental health situation, legal framework and recommendations about public policy on mental health. Its diagnosis reported similar challenges as in previously mentioned studies and NMHP analysis. However, it also underlined the relevance of the social environments such as work, school, household, and virtual. It recommended enhancing intersectoral coordination, strengthening healthy environments, and improving social inclusion for people with mental diseases, substance abuse, and victims of violence to address these challenges (36).

### Mental health definition

Mental health is presented as a holistic concept, described as actively influenced by sectors such as education, housing, work, culture, and sports, which are closely related to wellbeing and quality of life. The NMHP describes mental health as a historical and dynamic process, where, depending on the context, people’s actions, their relationships, and their potential are developed (2). As suggested by one of the stakeholders, ‘Historical’ is the word used in the reviewed documents to refer to the fact that the definition of mental health has evolved throughout history and is affected by historical events.

Mental health policy documents stated aims like achieving the development of wellbeing actions and enjoyment of a healthy life, promotion of healthy conditions and lifestyles where people, families and communities grow, work and age. In this sense, throughout documents there was an attempt to differentiate and respond to the mental health needs of each population and environment. The mention of “growing, working, and aging” makes reference to considering mental health throughout the life cycle.

### Mental health structures and actors involved

Institutional documents such as the NMHP outlined the different professionals carrying out mental healthcare, this included mental healthcare workers such as nurses, healthcare assistants, general practitioners, psychologists, and psychiatrist. Professionals’ work is directed to the promotion and prevention of mental health problems, and provision of integral healthcare in terms of diagnosis, treatment and rehabilitation. Furthermore, other documents such as the MinSalud guidelines on Attention to Mental Problems and Disorders within the framework of Peace Construction and Consolidation suggests including also social workers, sociologists, anthropologists, psycho-educators, artists, and local and community agents (29).

According to the NMHP, there are 11 mental health services that must be part of the mental healthcare provider’s plan:

1. Outpatient care.
2. Home care.
3. Pre-hospital care.
4. Care centres for drug addiction.
5. Community mental health centres.
6. Support groups for patients and families.
7. Adult day-hospital.
8. Children and young people day hospital.
9. Community-based rehabilitation.
10. Mental health units.
11. Psychiatric emergencies.

### Focus on early childhood and education

Family is presented as the main agent of social, economic, and cultural transformation. Strategies were centred around enhancing parenting by developing educational guidelines; strengthening mental health support in schools by monitoring violence (bullying) cases using the School Coexistence Information System (Sistema de Información Unificado de Convivencia Escolar in Spanish); and promoting key competencies outlined by WHO for children and young people, such as empathy, affective and assertive communication, or conflict resolution.

Institutions leading this work include the Ministries of Culture, Health, Justice, Housing, Sports, and the Colombian Institute for Family Welfare (ICBF in Spanish). They focus their efforts on promoting children’s and young people’s healthy development through interventions focusing on leisure time activities (i.e. extracurricular activities), increasing young people’s social participation by developing youth mental health networks and peer support groups, among others. Preventive actions included performing clinical screening tests to identify emerging mental health problems such as drug consumption and depression.

One stakeholder emphasised that substance abuse, including alcohol, tobacco, and vapers, should be the focus in terms of mental health in children and young people, however, the limited capacity of the mental health services in Colombia has led to an increased gap between care needs and provision. Others pointed out that the lack of implementation of “*these promising ideas*” for prevention in children and young people demonstrated the internal issues within public institutions, suggesting that a key aim should be to facilitate cross-institutional collaborations for implementation of mental health interventions. For example, institutions such as the Ministry of Culture and Ministry of Sports should be more involved in mental health intervention planning. They highlighted that factors such as violence, physical activity, and nutrition are undermined in the discussion of mental health in children and young people in practice.

### Culture and specific strategies for minorities

Culture is presented as a protective factor for mental health and involves the acknowledgement of others’ beliefs, values, and rituals. Accepting others’ point of view fosters interaction at the individual, family, and community levels. Documents from the Ministry of Health specifically dedicated to considerations around indigenous communities, highlighted the importance of culture. Emphasis was made on the significance of orientating mental health services towards intercultural approaches promoting worldviews, values, rituals and beliefs that favour interaction processes at the community, family and individual level. However, without disregarding attention to major clinical mental health priorities in these communities, such as mental health promotion, substance abuse, and suicide prevention. For this, documents referred to the use of training programmes like the WHO’s Mental Health Gap Action Programme (MhGAP) to characterise, canalise, intervene, and track the mental health needs of indigenous individuals. Due to the disperse geographical distribution of many of these communities, tele-mental health strategies were encouraged. Providers were recommended to provide healthcare staff with training on telemedicine, particularly supported by the Ministry of Communication and Technologies (MinTIC) to guarantee technology and internet access in all areas.

Additionally, the Department for Social Prosperity has increased awareness on mental health promotion topics in targeted communities such as immigrants, indigenous, and deprived communities, all of which aimed to overcome poverty, and social gaps and reach social equity.

There are also a series of documents dedicated specifically to mental health interventions in relation to the armed conflict. (37–44). For example, interventions for ex-combatants are recommended to be carried out through group and participatory methodologies to support the strengthening of coping capacities in the face of the changes and challenges involved in the process of re-incorporation into civilian life.

### Determinants of mental health, promotion, and prevention

All documents were directed to the development of actions that promote wellbeing and enjoyment of a healthy life. Mental health promotion included healthy conditions and lifestyles where people, families and communities grow, work and age. The latter part of this objective makes reference to the different stages of the life cycle which are all included.

A key focus is that of creating “resilient, healthy and protective environments”. A range of intra- and trans-sectoral actions were outlined for this purpose of strengthening environments and protective factors. For example, improving mobility and road safety within the framework of the Primary Health Care Strategy; creating favourable spaces for dialogue, conflict resolution, and network support; organising cultural meetings of community leaders that promote positive leadership; and prevention of suicidal behaviour and early detection during general medical care, gynaecology and paediatrics. Included within this were strategies specifically to prevent epilepsy, such as, managing maternal-perinatal health and prevention of road mobility accidents to avoid neurological and psychiatric sequelae; expanding the coverage of the Expanded Immunization Plan (PAI, in Spanish) and prevention against parasites such as cysticercosis to reduce infections of the central nervous system; and improving the control of chronic non-communicable diseases and cerebrovascular risk.

MinSalud established the essential determinants of mental health that should guide mental health interventions. Among them, social inclusion, stigma and discrimination eradication, good behaviour and violence prevention, school harassment prevention, suicide and drug use prevention, social involvement, and economic and food security. Intersectoral interventions to address them included *Familias en Acción* programme (conditioned cash-transfer scheme), intensive stigma and discrimination interventions on specific groups such as LGBTI, disabled, and homeless populations, among others.

Policies aimed to address mental health determinants, satisfying needs and reaching the highest level of quality of life in both individuals and communities. Multiple sectors participate in mental health promotion strategies following the Mental Health Law and National Mental Health Policy. For instance, MinTIC established guidelines regarding mental health communication in media. This provided the standards for broadcasting mental health-related content for all the country through both private and public media companies.

One stakeholder highlighted the importance of identifying the main social determinants of mental health in Colombia. The expert used as an example, how improving nutrition habits and physical activity behaviours leads to better mental health, but this was not visible nowadays in the mental health discussion in Colombia.

### Psychosocial Rehabilitation

Regarding psychosocial rehabilitation, the NMHL states the aim to “*facilitate the opportunities for individuals affected by a mental health illness to reach their maximum level of independent performance within their communities.*” To accomplish this objective, MinSalud established healthcare delivery routes (*RIAS,* in Spanish) focused on mental health promotion and care.

Older adults are repeatedly mentioned throughout rehabilitation documents. There is interest in “active ageing” based on developing support and aid groups, institutional networks, and social inclusion practices for seniors and caregivers. To accomplish this, strategies such as home visits are suggested to identify necessities, support families, and facilitate monitoring.

Community Based Rehabilitation (CBR) is described in the Ten-Year Public Health and National Mental Health Policy. CBR is an essential part of mental health and inclusion policies in Colombia. Institutions aim to guarantee the development of equal capacities for disabled individuals with an approach to prevention, care, and mitigation of events associated with mental health.

Adequate environments are presented as vital to facilitate inclusion and rehabilitation. Documents underlined the importance of protecting places where families and communities share, such as work, schools, households, universities, and neighbourhoods.

Yet, it is still necessary to adapt mental healthcare pathways to outpatient services, seeking to provide timely and appropriate care framed on risk management and cost reduction. Two stakeholders mentioned that CBR has not been implemented as it should be, since these services have been linked extremely to the health system and not to other non-clinical options. They stated this has led to an ‘institutionalised’ approach, which actually involve the community.

### Mental health policy assessment – WHO

We completed the WHO checklist for evaluating a mental health policy to detect content issues and evaluate overall performance (10).

Overall, of 76 questions/statements, 45 were accomplished to great degree, 13 were unknown, ten were not accomplished, and 7 were accomplished to some extent.

The NMHP is strengthened by being evidence-based on three different cross-sectional studies in the last 30 years. Also, the NMHP development was led by high-level mandate such as diverse ministries and governmental consulting bodies, with further discussions in the Congress. It also repeatedly addresses topics such as human rights, promotion, prevention, and rehabilitation.

However, the NMHP presented some limitations regarding the recommendations to consult diverse groups (i.e. not psychiatrists, but different mental health professionals, patient and family representatives, or NGOs). Moreover, there is no reported mention of information systems, advocacy, of quality improvement, representatives of relevant bodies such as Ministries of Finance or Housing.

Yet, as mentioned in the checklist instructions, specific roles from other relevant bodies, and more detailed information about MinSalud role might be described in parallel policy documents such as decrees, or executive orders. For instance, regarding coordination and management, the NMHP has a specific axe on this topic, but no detailed body or position within MinSalud is mentioned to oversee compliance. There is no major mention of financing in the document either. However, further information around this is provided in a parallel policy document (COMPES 3992). This is also true for some other items included in the WHO checklist which are described with more detail in documents other than the NMHP, such as the CONPES 3992 and the MHL.

The vision stated in the NMHP aims to prioritise mental health in the national agenda, considering previous related policies and laws. Four principles are mentioned: 1. Mental health as a key part of the right to health, 2. Health intercultural approach, 3. Social participation, and 4. Evidence-based public policy. The principles and axes emphasise and promote important topics such as human rights, intersectoral collaboration, community care, integration, among others. Though the vision, objectives and principles are consistent and articulated, the areas for action are written in a way that does not commit the Government (e.g., non-indication of action owner, use of should instead of will).

## Discussion

### Summary of findings

We conducted a review of policy, plan, and legislation documents currently in force to define mental health structures and understandings in Colombian policy. Most documents identified were created by the Ministry of Health, the Unit for the Attention and Integral Reparation to the Victims, and regional governments. We found Colombian policy had a holistic understanding of mental health, including considerations around the importance of prevention through the creation of healthy protective environments, as well as offering diagnosis, treatment and rehabilitation for people who require mental health care. There was a strong focus on childhood and addressing issues related to illegal drug consumption. Social determinants of mental health considered included social exclusion, violence, bullying, and economic instability. Numerous preventive programmes were listed such as specific programmes for LGTB+ and indigenous communities. These findings can be used as a starting point to map gaps between policy and practice, and policy and theoretical knowledge about mental health.

### Contrasting findings with existing literature

Like in most other countries, Colombian policy has evolved from disease-focused initiatives to health promotion in the last decade. This shift coincided with a time of heightened interest in the promotion of positive mental health and wellbeing, and increased evidence around the cost-effectiveness of prevention programmes (46, 47) that has continued being the global trend (48). Changing the focus has important implications for practice and future policy development, for example, an increase in psychosocial support interventions, community health education programmes, and community strengthening interventions. Effective programmes have been developed and tested for promoting mental health in everyday settings such as families, schools and workplaces. In a review conducted by our team in 2022, we found that out of twelve mental health interventions evaluated in Colombia, most were focused on treatment of substance use disorders and mental health disorders such as anxiety, depression, PTSD, and schizophrenia (49). While this review is not an exhaustive list of all the mental health interventions in the country, it hints to a focus still on ill mental health treatment in practice. However, many prevention and promotion interventions were highlighted in the policy documents, such as SACUDETE strategy for young people’s development, and the Casa Digna, Vida Digna programme.

As other modern public mental health policies, Colombian documents aim at improving psychosocial health by addressing determinants of mental health in all public policy areas. Research to identify key determinants of poor mental health in Colombia have consistently pointed to: limited schooling, relying on informal employment, exposure to violence and forced displacement, and most markedly, being a woman (50, 51). Colombian policy documents as a whole propose strategies that approach each of these social determinants, however it was noted that the essential determinants proposed by MinSalud to guide mental health intervention development do not coincide with this list. MinSalud’s proposed focus on topics such as social inclusion, good behaviour, and suicide and drug prevention may be more a list of critical topics based on the five axes of the NMHP, rather than evidence-based identification of health.

### Implication for policy, services and research

Research indicates that a significant number of mental disorders and suicides can be prevented through public mental health interventions. Available evidence supports a population-wide approach rather than focusing solely on high-risk individuals (52). Following the ‘Health in All Policies’ approach, mental health determinants should be addressed across various policy domains, extending beyond the health sector. It emphasizes the connections between mental health, productivity, quality of life, and in general with positive wellbeing outcomes (53). This approach serves as the foundation for many modern mental health policy documents, emphasising that promoting mental health effectively involves collaborating across various sectors, such as education, housing, employment, industry, transportation, arts, sports, urban planning, and justice, fostering a comprehensive approach. Optimal mental wellbeing is attained in societies that are fair, just, and free from violence. The most effective approach to promoting mental health involves inclusive and respectful methods that encourage participation, recognising and appreciating cultural heritage and diversity (54).

Future research needs to focus on understanding of resilience and formal and informal mental health delivery systems of care available in different Latin American countries (55).

### Strengths and Limitations

To our knowledge this is the first review of mental health policy documents in Colombia, thus filling an important gap in the literature. This review counts with important strengths due to methodological innovations applied to increase rigour and applicability of results in practice. These include a systematic approach to identifying, screening, and extracting information from policy documents; stakeholder consultations at every research stage to ground our study in real-world knowledge; and combining software and manual data analysis to reduce selection and confirmation bias, as well as have contextual sensitivity.

We should also acknowledge several limitations that impact the comprehensiveness of its findings. Firstly, the exclusion of mental health documents or guidelines from private institutions is a notable constraint, considering Colombia’s clinical mental health provision is still primarily carried out by private services. Consequently, the insights provided may not fully encapsulate the breadth of the mental health landscape, particularly within privately-operated services. We mitigated this bias by including the perspective of two stakeholders involved in private practice.

Additionally, despite efforts to incorporate diverse perspectives, challenges were encountered in ensuring the representation of all stakeholder groups from the start of the project. In particular, the valuable input provided at the later stages by user survivors, who suggested revisions to specific documents in future policy reviews, and an in-depth analysis of case laws. In relation to the latter, they deemed legal sentences from high courts, such as the “Fuente de Oro” case, “critically relevant” for understanding barriers to accessing mental health services. However, due to limitations in resources and scope, the paper does not delve extensively into these legal dimensions, leaving a potentially valuable aspect unexplored. Readers should approach the findings with an awareness of these limitations, recognising the need for further exploration and consideration of diverse viewpoints for a more comprehensive understanding of Colombian mental health policies.

## Conclusion

Results highlight Colombian mental health policy documents are comprehensive and for the most part up to date with international consensus for best practices in mental health. However, challenges remain in Colombia to (1) define roles and fostering patient and public involvement, (2) develop structures to produce local evidence for evidence-based planning, and (3) consolidate means to monitor the progress of policy implementation on the ground.

## Supporting information

Appendix 3 WHO checklist

## Data Availability

All data used in this study is available for public access through the institutional websites where they were identified.

## Acronyms

CBR: Community Based Rehabilitation.
CONPES: Consejo Nacional de Política Económica y Social in Spanish.
DANE: The National Administrative Department of Statistics / Departamento Administrativo Nacional de Estadísticas in Spanish.
DNP: National Planning Department / Departamento Nacional de Planeación in Spanish.
ICBF: The Colombian Family Welfare Institute / Instituto Colombiano de Bienestar Familiar in Spanish.
LGTB+: Lesbian, gay, bisexual, transgender, and queer people.
LISTEN: The Collaborative and Digital Analysis of Big Qual Data in Time Sensitive Contexts.
MhGAP: WHO’s Mental Health Gap Action Programme.
MHL: Mental Health Law / Ley Nacional de Salud Mental.
MINSALUD: Ministry of Health and Social Protection / Ministerio de Salud y Protección Social in Spanish.
MinTIC: Ministry of Communication and Technologies.
NGOs: non-governmental organisations.
NMHP: National Mental Health Policy / Política Nacional de Salud Mental.
OSF: Open Science Framework.
PDET: Programa de Desarrollo con Enfoque Territorial in Spanish.
PRISMA: Preferred Reporting Items for Systematic Reviews and Meta-Analyses.
RIAS: Rutas integrales de atención en salud in Spanish.
VPN: virtual private network.
WHO: World Health Organization.

