## Appendices 1 and 2 for "What do we have here? A Systematic Review of Mental Health Policy in Colombia"

### Appendixes

***Appendix 1.*** Search strategy

| Website/Organisation | Strategy |
|---|---|
| The National Department of Planning | salud mental AND PDET AND servicio salud mental AND presupuesto |
| Department of Social Prosperity | salud mental |
| Senate of Colombia and House of Representatives | |
| Local government of Florencia | |
| Local government of Caquetá | |
| The Victims Unit | |
| The Constitution of Colombia | Mental |
| | bienestar |
| The Ministry of Health | servicios AND salud mental |
| | salud mental" AND presupuesto |

***Appendix 2.*** Data extraction form

**Identification**

- Document title
- Search tool
- Publication date
- Type of document
- Organisation / Institution name ( Who issued the document)
- Population
- Type of services - interventions
- Reference / URL
- Date document was accessed

**Interest topics**

- Substance use
- Suicide
- Government / State support offered to people with mental health problems
- Human resources
- Budget - financing
- Level of attention
- Type of service
- Human rights
- Life course
- Gender
- Distinctions based on population / territorial characteristics
- Psychosocial

**SPICE model**

- Prosperity
- Individual recovery
- Social recovery
- Clinical recovery experience
