## Appendix 3 WHO checklist for "What do we have here? A Systematic Review of Mental Health Policy in Colombia"

### CHECKLIST FOR EVALUATING A MENTAL HEALTH POLICY

| Please use the following rating scale to rate each item:1 = yes/ to great degree 3 = no/not at all 2 = to some extent 4 = unknown | Rating | If "yes" or "to some extent"please state how. If not, please state reason(s). | Action required (if any) |
| --- | --- | --- | --- |
| **PROCESS ISSUES** | | | |
| 1a. Was there a high-level mandate to develop the policy (e.g. from the Ministerof Health)? | 1 | | |
| 1b. At what level has the policy been officially approved and adopted? (e.g., the department of mental health, Ministry of Health, Cabinet, Minister of Health). | 1 | It was approved at the national level and since then, it was officially adopted. | |
| 2. Is the policy based on relevant data: | | | |
| -- From a situation assessment? | 1 | Based on 3 descriptive cross-sectional studies in 1993, 2003, and 2005. | |
| -- From a needs assessment? | 1 | Based on 3 descriptive cross-sectional studies in 1993, 2003, and 2005. | |
| 3. Have policies relating to mental health that have been utilized within the country and in other countries with similar cultural and demographic patterns been examined and integrated where relevant? | 2 | Other local experiences were used as notable examples to guide implementation. Nevertheless, no international experience was considered. | |
| 4. Has a thorough consultation process taken place with the following groups: | | | |
| -- Representatives from the Health Sector, including planning, pharmaceutical , human resource development, child health, HIV/AIDS, epidemiology and surveillance, epidemic and disaster preparedness divisions. | 2 | Representatives from Colombia's psychiatry association. | It should have been more consultancy with other healthcare professionals' associations, pharma representatives, and epidemiology and surveillance institutions. |
| -- Representatives from the Finance Ministry? | 3 | It is not mentioned that Ministry of Finance participated in the policy. However, it participated in a parallel policy document (CONPES roadmap) | |
| -- Representatives from Social Welfare and Housing Ministries? | 3 | It is not mentioned that Ministry of Housing participated in the policy. However, it participated in a parallel policy document (CONPES roadmap) | |
| -- Representatives from the criminal justice system? | 1 | The Ministry of Justice participated through the guidelines to prevent to consumption of psychotropics. | |
| -- Consumers, or representatives of consumer groups? | 4 | | |
| -- Family members or their representatives? | 4 | | |
| -- Other NGOs? | 4 | | |

| | | | |
|---|---|---|---|
| -- Private sector? | 4 | | |
| -- Any other key stakeholder groups? If so, please list them | 4 | | |
| 5. Has an exchange taken place with other countries concerning their mentalhealth policies and experiences? | 4 | | |
| 6. Has relevant research been undertaken to inform policy development, (e.g.pilot studies)? | 1 | 3 descriptive cross-sectional studies in 1993, 2003, and 2005. Also, | |
| **CONTENT ISSUES** | | | |
| 1. Is there a realistic vision statement? | 1 | The vision aims to prioritise mental health in the national agenda, considering previous related policies and laws. | |
| 2. Are values and associated principles which inform the policy included? | 1 | The four principles are: 1. Mental health as a key part of the health right, 2. Health intercultural approach, 3. Social participation, and 4. Evidence-based public policy. | |
| 3. Do these values and associated principles emphasize and/or promote: | | | |
| -- Human rights? | 1 | Human rights are one of the principal approaches. | |
| -- Social inclusion? | 1 | Social inclusion is one of the main axes. | |
| -- Community care? | 1 | Community approach is mentioned multiple times | |
| -- Integration? | 1 | One of the axes focuses on articulation, integration, and coordination of different sectors. | |
| -- Evidence-based practice? | 1 | Evidence-based public policy is one key principles. | |
| -- Intersectoral collaboration? | 1 | One of the axes focuses on articulation, integration, and coordination of different sectors. | |
| -- Equity with physical health care? | 3 | It is not mentioned that mental and physical health should be equal. It is only mentioned that mental health is vital to have good physical and mental health. | |
| 4. Have clear objectives been defined? | 1 | | |
| 5. Are objectives consistent: | | | |
| -- With the vision? | 1 | | |
| -- With the values and principles? | 1 | | |
| 6. Are the areas for action clearly described to indicate the main policy directionsand what will be achieved? | 1 | Yes, they are divided in action axes, 5 in total. | |
| 7. Are the areas for action written in a way that commits the Government (e.g. dothey state "will" instead of "should")? | 2 | Document did not state neither will nor should. Document did not indicate action owner (e.g., enhance capacity without responsible) | It is important to establish responsible |

| | | | |
|---|---|---|---|
| | | | per action to reach accountability. |

| | | | |
|---|---|---|---|
| **8.** To what extent do the areas for action comprehensively address **coordination& management?** | | | |
| (a) Does the policy specify a dedicated mental health position/post within the Ministry of Health to coordinate mental health functions and services? | 4 | | The document has a specific axe on management and coordination. However, no detailed body or post is mentioned to oversee compliance. |
| (b) Does the policy establish or refer to a multisectoral coordinating body to oversee major decisions in mental health? | 4 | | |
| **9)** To what extent do the areas for action comprehensively address **financing?** | | | |
| (a) Does the policy indicate how funding will be utilized to promote equitable mental health services? | 4 | | There is no significant mention of financing in the document. However, further information is provided in a parallel policy document (COMPES roadmap) |
| (b) Does the policy state that equitable funding between mental health and physical health will be provided? | 4 | | |
| (c) If health insurance is utilized in the country, does the policy indicate whether/how mental health would be part of it? | 4 | | |
| **10.** To what degree do the areas for action comprehensively address **legislation and/or human rights?** | | | |
| (a) Does the policy promote human rights? | 1 | | |
| (b) Does the policy promote the development and implementation of human-rights-oriented legislation? | 1 | | |
| (c) Is the setting up of a review body envisaged to monitor different aspectsof human rights? | 1 | | |
| 11. To what extent do the areas for action comprehensively address **organization of services?** | | | |
| (a) Does the policy promote the integration of mental health services into general health services? | 1 | | |
| (b) Does the policy promote a community-oriented mental health approach? | 1 | | |
| (c) Does the policy promote deinstitutionalization? | 1 | | |

| | |
|---|---|
| 12. To what extent do the areas for action comprehensively address **promotion, prevention and rehabilitation?** Does the policy make provision for: | |
| (a) The prevention of mental disorders? | 1 |

| | | | |
|---|---|---|---|
| (b) Interventions that promote mental health? | 1 | | |
| (c) Interventions for the rehabilitation of people with mental disorders? | 1 | | |
| **13.** To what extent do the areas for action comprehensively address **advocacy?** | | | |
| (a) Is the policy supportive of consumers and family organizations? | 1 | The policy mentions the enhancement of networks of social support, for families, patients, and communities. | |
| (b) Is there emphasis on raising awareness of mental disorders and their effective treatment? | 4 | | There is not specific mention of raising awareness |
| (c) Does the policy promote advocacy on behalf of people with mentaldisorders? | 1 | | |
| 14. To what extent do the areas for action comprehensively address **quality improvement?** Does the policy | | | |
| (a) Make a commitment to providing high quality, evidence- based interventions? | 1 | The policy established one of its principles to be a public policy based on scientific evidence. | |
| (b) Include a process to measure and improve the quality of services? | 3 | | There is no mention detailed mention of improvement of quality of services |
| 15. To what extent do the areas for action comprehensively address **information systems?** | 2 | | It is mentioned, but only regarding integral care, which is one of the five axes. |
| (a) Will mental health information systems be set up to guide decision-making for future policy, planning and service development? | 3 | | No mention of further involvement of information systems in other areas/sections. |
| **16.** To what extent do the areas for action comprehensively address **human resources and training?** | | | |
| (a) Does the policy commit to putting in place suitable working conditions for mental health providers? | 3 | | There is no mention of strategies to improve recruitment and retention of mental health providers or detailed information about working conditions of mental health providers. |
| (b) Have appropriate management strategies been discussed to improve recruitment and retention of mental health providers? | 3 | | |
| (c) Are training in core competencies and skills seen as central to human resources development? | 2 | | To develop comprehensive care networks, the policy established one strategy to carry out the management |

| | | | |
|---|---|---|---|
| | | | processes necessary for the operation such as the continuous training and sufficiency of human resources. |
| 17. To what extent do the areas for action comprehensively address **researchand evaluation?** | | | |
| (a) Does the policy emphasize the need for research and evaluation ofservices and of the policy and strategic plan? | 1 | | |

| | | | |
|---|---|---|---|
| 18. To what extent do the areas for action comprehensively address **intrasectoral collaboration** within the health sector? Does the policy:: | | | |
| (a) Emphasize collaboration with planning, pharmaceutical, human resource development, child health, HIV/AIDS, epidemiology and surveillance, epidemic and disaster preparedness divisions, within the health sector? | 2 | | There is a specific |
| (b) Contain clear statements of what role each department will play in each area for action? | 3 | | No mention by might be included in related documentation. |
| 19. To what extent do the areas for action comprehensively address **intersectoral collaboration?** Does the policy: | | | |
| (a) Emphasize collaboration with all other relevant government departments? | 1 | | |
| (b) Emphasize collaboration with all relevant NGOs, including consumer and family groups? | 1 | | |
| (c) Contain clear statements of what role each sector will play in each areafor action? | 4 | | |
| 20. Have all of the following groups been considered: | | | |
| -- People with severe mental disorders? | 1 | | |
| -- Children and adolescents? | 1 | | |
| -- Older persons? | 1 | | |
| -- People with intellectual disability? | 2 | | People with intellectual disability are grouped within the disabilities categories, but no specific details are provided. |
| -- People with substance dependence? | 1 | | |
| -- People with common mental disorders? | 1 | | |
| -- People affected by trauma? | 1 | | |
| 21. Given resources available in the country, has a reasonable balance been achieved between the above groups? | 1 | | |

| | |
|---|---|
| 22. To what degree have the key mental health policy issues been integratedwith/or are consistent with the country's: | |
| -- Mental health law? | 1<br>1111 |

| | |
|---|---|
| -- General health law? | 1 |
| -- Patients rights charter? | 1 |
| -- Disability law? | 1 |
| -- Health policy? | 1 |
| -- Social welfare policy? | 1 |
| -- Poverty reduction policy? | 3 |
| -- Development policy? | 3 |

**Taking into account the financial and human resources available in the country, comment on the general feasibility for implementation of the policy.**
